# EXPLORING HIGH MORTALITY RATES AMONG PEOPLE WITH MULTIPLE AND COMPLEX NEEDS: A QUALITATIVE STUDY USING PEER RESEARCH METHODS

**DOI:** 10.1101/2020.11.23.20235416

**Authors:** Rachel Perry, Emma A. Adams, Jill Harland, Angela Broadbridge, Emma L. Giles, Grant J. McGeechan, Amy O’Donnell, Sheena E. Ramsay

**Affiliations:** Population Health Sciences Institute, Newcastle University, Newcastle upon Tyne, United Kingdom; Fulfilling Lives Newcastle Gateshead, Gateshead, United Kingdom; School of Health and Life Sciences, Teesside University, Middlesbrough, United Kingdom; Centre for Applied Psychological Science, Teesside University, Middlesbrough, United Kingdom

**Keywords:** multiple complex needs, mortality, prevention, interventions

## Abstract

**Objective:** To explore the reasons underlying high mortality rates among people with multiple and complex needs and potential preventive opportunities.

**Design:** Qualitative study using peer research

**Setting:** North East of England

**Participants:** Three focus group discussions were held involving: 1) people with lived experience of MCN (n=5); 2) frontline staff from health, social care, and voluntary organisations that support MCN groups (n=7); and 3) managers and commissioners of these organisations (n=9).

**Results:** Findings from this study provide valuable insights from people with lived experience and staff on the complexity underpinning high mortality rates for individuals experiencing multiple and complex needs. Mental ill-health and substance misuse (often co-occurring dual diagnosis) were highlighted as significant factors underlying premature mortality among multiple and complex needs groups. Potential triggers to identify people at-risk included critical life events (e.g. bereavement, relationship breakdown) and transitions (e.g. release from prison, completion of drug treatment). Early prevention, particularly supporting young people experiencing adverse childhood experiences was also highlighted as a priority.

**Conclusion:** High mortality in MCN groups may be reduced by addressing dual diagnosis, providing more support at critical life events, and investing in early prevention efforts. Future service delivery should take into consideration the intricate nature of multiple and complex needs and improve service access and navigation.

**ARTICLE SUMMARY:** *Strengths and limitations of this study:*

- This study employed focus group discussions with individuals with multiple and complex needs and service providers to understand the complexity underpinning high morality rates for individuals experiencing multiple and complex needs.
- Peer researchers contributed to all stages of this study, including developing the aims, data collection, interpretation, and shaping recommendations.
- Using peer researchers enhanced our access to participants and improved interpretation of data
- The main limitation is that the study only recruited individuals in one region in the North East of England. Views from individuals with MCN and service providers in other areas of England might have led the results to being more generalisable.

## INTRODUCTION

Despite increases in life expectancy in the United Kingdom (UK) throughout the 20^th^ century, improvements in mortality rates have stalled in recent years for most population groups.^1^ Since 2010-2011, the gap in life expectancy between people living in the most and least deprived communities has widened, with the largest decreases seen in the 10% most deprived neighbourhoods in the North East of England. ^2, 3^ This region has also observed high rates of drug-related deaths and suicide in younger adults (under age 50) compared to other parts of the UK.^4^

People facing co-occurring issues of homelessness, substance misuse, offending, and mental ill-health experience particularly poor health outcomes compared to the general population.^5^ Multiple complex needs (MCN) has been used to describe this group; other terminology include severe and multiple disadvantage or multiple exclusion homelessness.^5-8^ When considered in isolation, each of these issues contributes to higher mortality rates compared to the general population.^9-12^ However, there is a growing appreciation of the intersecting nature of these needs and the compounding effect these can have on health outcomes.^5^ All-cause mortality rates for people with MCN compared to the UK general population is almost seven times higher for men and almost twelve times higher for women, with major causes of deaths including injury, poisoning, external causes (i.e. accidents, homicide, or suicide), and social exclusion.^13^

Despite such inequality, gaps remain in our understanding of the perspectives of individuals with lived experienced of MCN and those who support them (e.g. health and social care workers) on the factors contributing to high death rates in this population.^14^ Involving peer researchers across the continuum of research, from concept development, to data collection, analysis, and dissemination, can help provide valuable access to participants from marginalized populations, as well as offering unique insights on how evidence should be collected and interpreted.^8, 15-17^ Peer researchers are individuals who have lived experience of the phenomenon under study and are at a stage where they are ready to support others discuss their own experiences.^8^ Although there is increasing recognition of the need to involve peer researchers in health and social care research, there remains limited representation in current literature on mortality and MCN.

The aim of this study was to explore the perspectives of people with lived experience of MCN and professionals who support them in order to understand factors underlying high mortality rates in this population. In doing so, we also sought to obtain rich insights into opportunities for preventing deaths and improving service provision for MCN groups in the future. The study was developed and undertaken with peer researchers.

## METHODS

A qualitative study using a peer-research approach was conducted. Peer researchers from Fulfilling Lives Newcastle Gateshead, a project that supports MCN groups, contributed to all stages of this study, including developing the aims, data collection, interpretation, and shaping recommendations. Peer researchers received NVQ-level training in peer research and had lived experience of MCN.

### Data collection

The study sample comprised people with lived experience of MCN, frontline staff, and managers/commissioners of relevant health, social care, and voluntary support services. Peer researchers suggested using focus group discussions (FGDs) to help create a supportive environment for conversations around a sensitive topic. A combination of convenience (using pre-existing networks) and maximum variation sampling techniques were used to recruit participants through email and mailing lists within the North East of England for three homogenous FGDs. Participants were recruited from both operational (frontline) and commissioning/managerial levels to ensure diversity of staff perspectives. Additionally, variation was sought in terms of gender, different components of MCN, and age for participants with lived experience, although all were aged 18 and over. Many peer researchers had relationships with lived experience FGD participants; however, this was a deliberate strategy as it enhanced our access to participants and improved the interpretation of data.

Ethical approval for the study was obtained from Newcastle University Ethics Committee (reference 11064/2018). A participant information sheet was made available to all participants and informed consent was gained prior to data collection.

Three homogenous FGDs were conducted in July 2019 with: individuals with lived experience of MCN (FGD 1, n=5); frontline staff (FGD 2, n=7); and managers/commissioners (FGD 3, n=9) working in health, social care, and voluntary sectors. Representatives from a range of voluntary and statutory organisations participated, including those working in local authority commissioning, mental health, substance misuse and housing and family services. No participants withdrew during the FGDs.

The FGDs were jointly facilitated by an academic and a peer researcher and held at accessible venues in Newcastle upon Tyne. Two semi-structured topic guides (Table 1) were co-produced with peer researchers, one for use with individuals with MCN and one for staff FGDs, with probes used to elicit additional information and detailed responses. All participants were fully briefed on the study (aims, objectives, and dissemination plans) prior to gaining consent. FGDs lasted approximately 90 minutes, and were audio recorded.

**Table 1.** **Topic guides for the focus groups**

| **Topic guide for focus groups with individuals with lived experience of MCN** | **Topic guide for focus groups with staff working with MCN groups** |
| --- | --- |
| • Awareness of mortality within their peer group<br>• What factors/life experiences do they think contribute to premature mortality within their peer group.<br>• Any concerns they have about this personally or for others<br>• Do they think anything could have been done to prevent people dying?<br>• Can they describe this? | • Awareness of premature mortality within multiple and complex needs groups<br>• Awareness of risk factors for premature mortality<br>• Current approaches to identify those at risk – perceptions of effectiveness<br>• What would help the services identify/ target those at risk<br>• Current interventions – perceptions of effectiveness |
| • What types of help and support would they like to see being developed/ provided?<br>• How would this be best offered? | • Types of interventions/approaches they think should be in place<br>• How could this be taken forward |

### Data analysis

Audio-recordings were transcribed verbatim and uploaded into NVivo 12 to support data management and analysis. To protect anonymity, particularly given the sensitivity of the topic, any potential identifiers (including participant characteristics such as age, gender, role) were removed. Field notes were used to capture immediate reflections on the FGDs and to aid analysis and interpretation. Peer researchers reviewed transcripts for authenticity. Deductive thematic analysis based on Braun and Clarke’s^18^ approach was conducted. Initial codes were co-created with peer researchers based on the study aim and objectives, existing literature, and an initial review of the transcripts. Each transcript was coded whilst adaptively developing the code book with any new codes and returning to previous transcripts. The first author conducted the primary coding and a section of each transcript was second coded by two other authors to ensure consistency. Codes were collated into potential themes, which were reviewed amongst the research team and further refined. In addition to peer researchers, the team compromised academic researchers, service providers, and public health registrar trainees with prior research experience. Consensus was reached through discussions with peer researchers and the wider research team on the themes, subthemes, extracts, and recommendations.

### Patient and public involvement statement

One of the co-authors (AB) works for Fulfilling Lives Newcastle Gateshead, which is an eight-year learning programm to improve the lives of people with MCN and build a trauma-informed approach with the services that support them. The project design and methodology was developed with a group of Experts by Experience (peer researchers) and fronline staff who work for Fulfilling Lives Newcastle Gateshead. As previously mentioned, peer researchers contributed to all stages of the research. The methodology selected for data collection (FGDs) was choosen based on suggestions from Peer Researchers.

## RESULTS

Findings were relatively homogenous across all three FGDs. Findings are presented according to the two main study objectives and associated themes: *understanding factors underlying premature mortality* amongst MCN groups and *opportunities for interventions to prevent mortality*.

### Understanding factors underlying premature mortality in MCN groups

#### Burden of mental ill-health and substance misuse issues

The severe burden of mental ill-health in people with MCN was a recurrent concern and felt to make a substantial contribution to premature mortality, particularly in relation to mental illness alongside substance misuse (dual diagnosis).

> *“Most of the people I know that’s died, their mental health has just been shot to bits, it’s all about the drugs. They’re taking the drugs because of mental health, is that bad? I’d say more the mental health killed them…the drugs just done that job”*
>
> -*Individual with lived experience of MCN*

There was a perception that people with MCN experienced challenges when trying to access services and support for co-existing mental ill health and substance misuse. Services were set up to deal with either mental health issues, or addiction issues, meaning those with a dual diagnosis failed to receive appropriate support.

> *“I just find that people who have got mental-health issues and also have addiction problems fall through the gaps, time and time again”*
>
> -*Frontline staff*

#### Lack of hope, stigma and health-seeking behaviour

Participants with MCN highlighted that experiences of loss were so common that it led to individuals becoming desensitized to deaths and reaching a point of acceptance. Outcomes for individuals experiencing MCN were so often negative this led to a lack of hope that it could ever be any different.

> *“You don’t see another way…it’s just doom and gloom and like you say this one’s dead, this one’s in prison, there’s nothing ever…it’s like being in the sort of devil’s dungeon, to be honest”*
>
> *-Individual with lived experience of MCN*

Stigma was highlighted as playing a large role in health-seeking behaviour. Individuals with lived experience expressed frustration around the stigmatizing nature of the words professionals used to describe different facets of MCN.

> *“I think that word as well, like, junky really boils my blood, heroin addict is much better. Just picking up the junky, junky, junky, that’s all we get*.*”*
>
> *-Individual with lived experience of MCN*

This fear of stigma along with not being given sufficient time for staff to understand their unique experiences led to individuals avoiding services. FGDs with staff reiterated this perception that many young people they worked with felt intimidated when going to see their General Practitioner and instead chose to self-medicate with illicit substances.

> *“… this doctor at the time of [the] appointment isn’t going to be able to comprehend even a tiny touch of what your life is*.*”*
>
> -*Individual with lived experience of MCN*
>
> *“We talked to the young people who have got mental health issues and they’re kind of like, “Oh no, I don’t want to talk to anyone, I don’t want to tell them I’ve got a problem. I’d rather just smoke some weed or take some grass and I’ll be okay*.*”*

*-Manager/Commissioner*

#### Poor navigation and limited services

Across all three FGDs, participants expressed problems with navigating service pathways, limited services and having to engage with traditional models of care. Individuals with MCN felt as though they were faced with a series of barriers to gaining appropriate support, which increased the vulnerability of MCN groups to premature mortality.

> *“I’m facing this maze full of doors and every time I open a door, there’s another door, sets of doors. There’s no coherent structure within the system that says, “Here’s a person who is asking for help, who’s engaging with everything that we’re giving, can we please pull this together so we can actually provide the help that this person needs*.*”*
>
> -*Individual with lived experience of MCN*

There was a perception that recent reductions in available support services meant that the support was often only provided in extreme circumstances, such as when an individual had reached a crisis point.

> *“it’s often such a desperate situation that we’re having ridiculous conversations that we want someone to be sectioned or we want someone to go to prison just so they’re in some kind of contained environment where we feel we can try and manage some of the risks”*
>
> -*Manager/Commissioner*

### Opportunities for interventions to prevent morality in MCN groups

#### Intervention timing

Critical life events (e.g. childhood adversity and bereavement) and transitions (e.g. release from prison) were highlighted as moments of key vulnerability, as well as potential opportunities for intervention. Participants felt that many of the issues encountered by people experiencing MCN were rooted in early childhood. Therefore, improved support for young people who experience adverse childhood experiences could prevent the development and exacerbation of long-term needs.

> *“Preventive measures early on may stop the numbers of people coming through with multiple and complex needs. So it’s the preventive, it’s the community centres, it’s the youth centres, it’s those things where the learning happens*.*”*
>
> *-Manager/Commissioner*

Participants in all focus groups agreed that windows of opportunity (time points of places where services could be provided) are often brief and difficult to take advantage of.

> *“There’s often an inability to exploit windows of opportunity where…support workers will try and get all their ducks in a row. So the mental health stuff, the mental health treatment, housing, benefits, all of that sort of stuff, it’s rare that you’re going to manage to get all of that sorted in the two hours of window opportunity you’ve got. Then the ship sails sometimes and you don’t know whether that’s going to come back again or when it’s going to come back again*.*”*
>
> *-Manager/commissioner*

#### Intervention approaches

Participants provided suggestions for how interventions could be improved in future. The need for a holistic, person-centred approach was highlighted, acknowledging that a “one-size-fits-all” approach was unlikely to cover the complexity of need or empower individuals to engage.

> *“I think there needs to be focus on it being really a person-centred approach and say, “This isn’t working for me at the moment and that’s how I would like things to be,” and giving them that sense of responsibility”*
>
> -*Frontline staff*

Building a sense of community for people living with MCN was a common suggestion aimed at reducing social exclusion. The value of peer support communities in particular was highlighted.

> *“There’s a connection [as a peer supporter]. Immediately there’s a connection but through that connection you feel like you’ve gotten in and you feel like what you say has a better chance of making a difference to that person”*
>
> -*Individual with lived experience of MCN*

The need for better collaboration across the multiple agencies involved in supporting people living with MCN was also highlighted. In particular, improved communication between services, and the opportunity to share learning across sectors were suggested as ways to reduce missed opportunities for prevention.

> *“We exist in a competitive tendering landscape and we need to leave that aside and come together and share good practice and learn from what’s happening across the world”*
>
> -*Manager/Commissioner*

## DISCUSSION

This study used FGDs to gather insights from individuals with lived experience, frontline workers and those responsible for managing and commission services on factors underlying high mortality rates in MCN populations and opportunities for potential interventions. Across discussions, we found that issues relating to the burden of mental ill-health and substance misuse (dual diagnosis), combined with experiencing stigma and exclusion when accessing services, were felt to contribute to adverse mortality outcomes in this group. Participants highlighted key life events (e.g. childhood adversity) and transitions (e.g. release from prison), as potential opportunities for targeted intervention to better support this population in the future.

Epidemiological data confirms the contribution of mental ill-health and co-occurring substance misuse to the high mortality rates in people experiencing MCN.^12, 19^ Homeless individuals have greater prevalence of alcohol use disorders than the general population,^20^ and self-reported data suggest that mental health problems are as high as 92% among individuals with MCN.^5^ This study provides valuable qualitative insights from both people living with MCN and those delivering and commissioning services as to how these issues combine to affect health outcomes. As others have reported, we found that the reality of many individuals with MCN is a journey dominated by the challenge of navigating a siloed, highly fragmented system, that is ill-equipped to meet their needs.^21^ Not unique to this study, are the experiences of many people with MCN finding the co-occurring nature of issues leads to an inability to access services or falling through system “cracks.”^22^ However, we also highlight the stigma and discrimination experienced by people with MCN as they attempt to access professional services designed to provide help, which represents a further barrier to support, as well as contributing to a deep sense of hopelessness amongst this population.

A recent mapping exercise of MCN across England found the current system of support across public and voluntary services was struggling to meet present demands and deliver positive outcomes for individuals with MCN.^5^ This is consistent with experiences shared throughout our study. Participants in the focus groups pointed to issues with the present system and identified that early prevention and targeting interventions at “critical life events” (i.e. transitions in service provision or at experiences of heightened adversity) could be important in reducing deaths in MCN groups. There was a particular emphasis on creating interventions targeting youth to address concerns earlier on in the prevention spectrum. Evidence highlighting the link between experiences of childhood adversity and later life MCN reiterate the need to create preventative interventions surrounding this time period.^5, 7, 23^ Additionally, our findings suggest that future provision should focus on interventions that are developed collaboratively across sectors ^5, 24^ targeting critical life events ^7, 23^ using person-centred and trauma-informed practices,^24, 25^ and peer support.^26^

The novelty of this study is it has shown the value of listening to individuals with experience of MCN. It enabled an exploration of an issue that directly affects their community and supported understanding of some of the underlying issues, as well as identifying some avenues for possible preventive interventions. This enrichment of the research aligns with existing literature exploring peer research use for vulnerable populations or sensitive topics.^8, 15-17^ Furthermore, the insights specific to opportunities for service provision take into consideration the lived experience of the target population, which can lead to more equitable service delivery and engagement.^16^

## Limitations

As our study was located in the North East of England, our findings may not be generalisable to other regions. Response bias is a possibility as individuals with stronger views might have been more likely to volunteer participate in a study. However, since focus groups were used, we feel that the potential effects were reduced as the groups allowed for a range of views to be considered. There were limitations in the representativeness of individuals with MCN; a recruitment challenge experienced in previous studies.^16^ Nonetheless, by engaging peer researchers throughout the research process, we feel this potential limitation was reduced. There is also the risk that participants might have felt more comfortable speaking to an outsider rather than a peer about certain issues.^17^ However, as the study focused less on the individual and more on the overarching issues related to mortality rates, this was likely minimal.

## CONCLUSIONS

Mortality rates among individuals experiencing MCN in the UK continue to rise due to an interconnected web of disadvantage and exclusion. Findings from this study provide valuable insights from people with lived experience and staff supporting MCN groups on the complexity underpinning this trend. Reducing mortality rates among MCN groups requires addressing issues related to dual diagnosis, providing support around critical life events, and investing in early prevention. The intersecting nature of MCN should be factored into the design of future interventions to address the challenges related to service navigation and availability. The need to address the underlying causes of MCN and develop effective and sustainable interventions to address high mortality rates has become even more important in light of the COVID-19 pandemic, which has exacerbated issues around service access, health inequalities, and social isolation.^27^

## Data Availability

Due to the sensitivity of the topic and the numbers of participants, we are not able to share FGD transcripts. Summaries are available from the corresponding author on request.

## Acknowledgements

We would like to acknowledge the various Peer Researchers from Fulfilling Lives Newcastle Gateshead who contributed to all stages of this study including developing the aim, methods, data collection, and interpretation. As well, we appreciate the time and effort of all participants who offered their views on and experiences of this challenging and emotive topic.

## Author contributions

Conceptualization: JH, SR, AOD, AB, GJM, ELG and RP

Data acquisition: JH, AB, and SR

Analysis: RP, AB, ELG, GJM, and EAA

Interpretation: All authors contributed to the interpretation of findings.

Writing: All authors contributed to the writing of the paper.

All authors have read and approved the final version to be submitted for consideration for publication.

